# REAL-TIME MECHANISTIC BAYESIAN FORECASTS OF COVID-19 MORTALITY

**DOI:** 10.1101/2020.12.22.20248736

**Authors:** Graham C. Gibson, Nicholas G. Reich, Daniel Sheldon

**Affiliations:** University of Massachusetts Amherst

## Abstract

The COVID-19 pandemic emerged in late December 2019. In the first six months of the global outbreak, the US reported more cases and deaths than any other country in the world. Effective modeling of the course of the pandemic can help assist with public health resource planning, intervention efforts, and vaccine clinical trials. However, building applied forecasting models presents unique challenges during a pandemic. First, case data available to models in real-time represent a non-stationary fraction of the true case incidence due to changes in available diagnostic tests and test-seeking behavior. Second, interventions varied across time and geography leading to large changes in transmissibility over the course of the pandemic. We propose a mechanistic Bayesian model (MechBayes) that builds upon the classic compartmental susceptible-exposed-infected-recovered (SEIR) model to operationalize COVID-19 forecasting in real time. This framework includes non-parametric modeling of varying transmission rates, non-parametric modeling of case and death discrepancies due to testing and reporting issues, and a joint observation likelihood on new case counts and new deaths; it is implemented in a probabilistic programming language to automate the use of Bayesian reasoning for quantifying uncertainty in probabilistic forecasts. The model has been used to submit forecasts to the US Centers for Disease Control, through the COVID-19 Forecast Hub. We examine the performance relative to a baseline model as well as alternate models submitted to the Forecast Hub. Additionally, we include an ablation test of our extensions to the classic SEIR model. We demonstrate a significant gain in both point and probabilistic forecast scoring measures using MechBayes when compared to a baseline model and show that MechBayes ranks as one of the top 2 models out of 10 submitted to the COVID-19 Forecast Hub. Finally, we demonstrate that MechBayes performs significantly better than the classical SEIR model.

## 1. Introduction

The emergence of COVID-19 in early 2020 led to the largest pandemic in over a century. Understanding the future trajectory of the pandemic can help decision-makers prepare for and consequently diminish the impact in terms of healthcare and economic burden. Forecasts of incident and cumulative deaths due to COVID-19 help in resource allocation and re-opening strategies [1]. Forecasts provide important data to decision-makers and the general public and can improve situational awareness of current trends and how they will likely evolve in coming weeks.

Infectious disease forecasting, at the time horizon of up to 4 weeks in the future, has benefited public health decision makers during annual influenza outbreaks [2, 3]. However, many forecasts of endemic, seasonal diseases, such as influenza, rely on ample historical data to look for patterns that can be projected forward into the future. In an emerging pandemic situation, models must be able to fit to limited data.

With limited historical data, mechanistic models are a natural framework for modeling and forecasting COVID-19. These directly model the transmission of the disease through the population and can be fit to public health surveillance data with relatively few parameters.

Our work is based on compartmental models, which are classical mechanistic models for disease transmission that were first introduced by Kermack and McKendrick [4]. These assume that, at any given time, each individual is in one of a mutually exclusive set of compartments, typically either the susceptible, exposed, infected, or recovered compartment. A model is specified by setting the rates of flow of individuals between compartments. While these models have been used since their inception in the early 20th century, the COVID-19 pandemic represents a unique opportunity to explore their forecasting properties in real-time at local, national, and global scales, for an emerging pathogen that, unlike influenza, does not have years of data on which models can train.

We develop a mechanstic Bayesian model (“MechBayes”) that tailors compartmental models to the operational needs of making one- to four-week ahead forecasts of incident deaths for COVID-19. Since the primary goal is to parsimoniously forecast an observable quantity, identifying internal parameters of the model, many of which are poorly determined or not identifiable from the available data [5], is not an explicit focus or prerequisite of our work. We distinguish this set-up from longer-term scenario projection models, which require well identified epidemiological parameters that can be set to counterfactual values under different scenarios, such as an increase or decrease in intervention levels [6, 7]. Scenario projection models are often based on similar foundations, but require different adaptations than those needed for real-time forecasting.

MechBayes is tailored to the particular needs and data availability of COVID-19. The compartmental model jointly models infections and deaths and uses records of both incident cases and deaths—the two most widely available COVID-19 surveillance measurements— for model fitting. MechBayes includes components to model changes over time in both the dynamics and the detection of COVID-19. In particular, transmission rates have changed significantly due to the addition and removal of control measures such as social distancing, lockdown, and mask use, while infection reporting rates have changed due to significant changes in the availability of diagnostic testing.

Recent forecasting efforts have recognized the need for probabilistic forecasting, with statements about uncertainty of the forecast relaying important public health information [8]. We adopt a Bayesian approach, which is naturally suited to quantifying uncertainty in parameters and forecasted quantities. MechBayes is implemeted in the NumPyro probabilistic programming language [9], which automates the complex task of designing a posterior sampling algorithm. NumPyro uses the JAX Python library [10] to automatically compute the partial derivatives needed for sampling via Hamiltonian Monte Carlo [11], and to compile model code for highly efficient computations. JAX includes a differentiable solver for ordinary differential equations (ODEs) [12], which allows us to embed ODE-based compartmental models into the full probabilistic model with relative ease. We are not aware of prior work that use numerical simulation of ODE-based compartmental models within a fully Bayesian framework, though there are other approaches to adapt compartmental models for Bayesian analysis [13]. The ability to build a modular probabilistic model with complex components and automatically obtain efficient inference procedures is a testament to recent advances in algorithms and software packages largely driven by applications in artificial intelligence and deep learning.

We demonstrate the success of the MechBayes by showing that forecasts submitted to the US Centers for Disease Control via the COVID-19 Forecast Hub outperform a baseline probabilistic forecast model [1, 14]. We additionally show that MechBayes is one of the top performing models out of those submitted to the COVID-19 Forecast Hub. Finally, we quantify the important features of MechBayes via an ablation study.

## 2. Methods

### 2.1. Data

In this analysis we used confirmed case counts and deaths for the 50 US states and the District of Columbia as reported by the Johns Hopkins University Center for Systems Science and Engineering (JHU CSSE) [15]. The data set reports the incident number of confirmed cases and deaths for each location at a daily frequency starting in early 2020. As noted in [16], COVID-19 cases are under-reported, with the fraction of all infections reported as cases for the US estimated at 20-30% [17]. The fraction of all infections reported has also changed dramatically over the course of the epidemic [18].

### 2.2. Forecast Targets

We made probabilistic forecasts for 1–4 week ahead incident and cumulative deaths for all geographies. An individual forecast distribution is represented by a set of 23 quantiles, ℚ = {.01, .05, .10, …, .90, .95, .99}, with the median (.50 quantile) representing the point forecast. While forecasts are made at the daily scale, we aggregate them to the weekly scale by summing incident death forecasts from the first forecasted Sunday through the following Saturday. We evaluate only forecasts of incident deaths, which is the primary modeled quantity; forecasts for cumulative deaths are created by accumulating forecasted incident deaths.

### 2.3. Mechanistic Bayesian Model

Compartmental models have been used to effectively model and forecast disease in non-pandemic situations both retrospectively and in real-time. These include complex compartmental models for real-time influenza forecasting [19, 20, 21], and a retrospective model evaluation of the 1918 influenza pandemic [22]. Compartmental models have been used for both inference and forecasting not just in respiratory disease but in Ebola [23], measles [24], dengue [25] and a wide variety of other communicable diseases.

Compartmental models have also been adopted into a Bayesian framework before, including both stochastic disease dynamics and deterministic dynamics [26, 27]. Non-parametric transmissibility was included in a Bayesian SEIR model to study Ebola by Frasso and Lambert [13]. Time-varying transmissibility has also been studied in the frequentist setting using complex non-parametric functions [28]. Many efforts have been made to use SEIR models in forecasting COVID-19 [29, 30, 31, 32, 33]. With the outbreak of COVID-19, accounting for testing has become a critical element in effectively using an SEIR model [6, 34].

The MechBayes probabilistic model consists of three parts, which together define a probabilistic model for the observed incident cases and deaths with the parameters and state variables of a compartmental model as latent variables. The core part is the mechanistic disease model *p*(*x*_1:*T*_, *η*_1:*T*_ |*θ*), which defines the distribution of the state variables *x*_1:*T*_ and time-varying parameters *η*_1:*T*_ given a vector *θ* of fixed, non–time-varying, parameters. The state variable *x*_*t*_ is a vector that enumerates the number of individuals in each compartment (susceptible, exposed, infectious, etc.) at time *t*, while *η*_*t*_ contains parameters of the disease model or observation process that change over time (e.g., due to changes in social distancing or test availability), which we model stochastically. MechBayes operates at a daily time step. The state trajectory *x*_1:*T*_ = (*x*_1_, …, *x*_*T*_) concatenates state vectors from each day, and *η*_1:*T*_ collects time-varying parameters in a similar fashion. MechBayes also defines a prior distribution *p*(*θ*) over fixed parameters, and an observation model *p*(*y*_*t*_|*x*_*t*_, *θ*) for the vector *y*_*t*_ of observed variables at time *t* (incident cases and deaths) given the state vector *x*_*t*_, time-varying parameters *η*_*t*_, and fixed parameters *θ*. Each part of the probabilistic model is expressed by writing Python code to sample from the corresponding distribution within the NumPyro probabilstic programming language [35], which automates the construction of Markov chain Monte Carlo algorithms to sample from the distributions *p*(*θ, x*_1:*T*_, *η*_1:*T*_ |*y*_1:*T*_), for inference about unobserved parameters and state variables, and *p*(*y*_*T* +1:*T* +*k*_|*y*_1:*T*_), for forecasting (by integrating over unobserved state variables and parameters).

#### 2.3.1. Disease Model

The MechBayes compartmental model is illustrated in Figure 1 and is based on the classical SEIRD framework [5]. It consists of state variables *S, E, I, R, D*_1_, *D*_2_ that indicate the number of individuals in the population that belong on a given day to each one of the following mutually exclusive compartments: susceptible (*S*), exposed (but not yet infectious) (*E*), infectious (*I*), recovered (*R*), or one of two death compartments (*D*_1_ and *D*_2_).^1^ The death pathway is separated into two compartments to incorporate a time-delay between infection and death that is modeled separately from the ratio between observed cases and observed deaths, which both have prior estimates from the literature [36]. For simplicity, we assume a closed population of size *N*. The following parameters govern how members of the population move between compartments:

- *β*_*t*_: transmission rate, which we allow to vary by time *t*;
- *σ*: rate of transition from the exposed state *E* to infectious state *I*; i.e., 1*/σ* is the expected duration of the time between exposure and onset of infectiousness;
- *γ*: rate of transition from the infectious state *I* to either *D*_1_ or *R*; i.e., 1*/γ* is the expected duration of the infectious period;
- *ρ*: fatality rate (i.e., probability of transitioning from *I* to *D*_1_ as opposed to *R*);
- *λ*: rate of transition from *D*_1_ to *D*_2_ (i.e., the inverse of expected number of days in *D*_1_ compartment before death)

**Figure 1.**
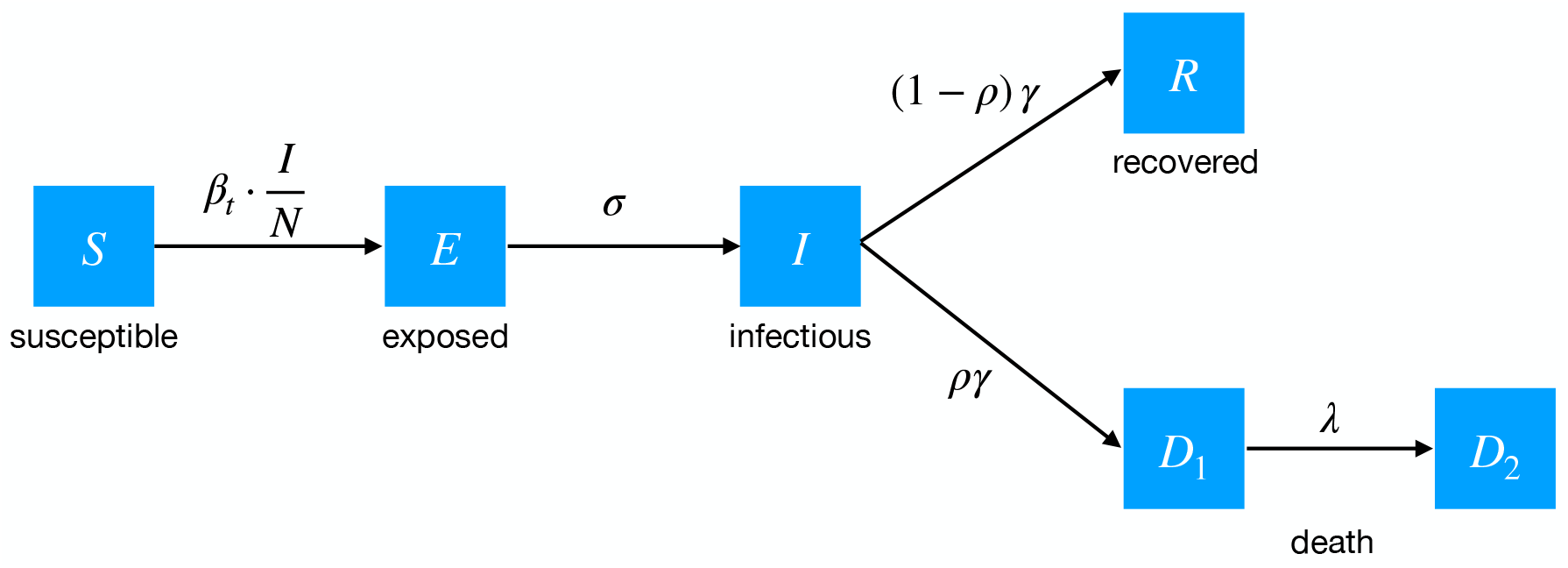
Flow diagram for MechBayes. Susceptibles (*S*) become exposed (*E*) with a rate of 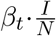 (proportional to the number of infected and infection probability times average number of contacts). Exposed individuals become infections with a mean time of 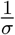. Infectious individuals can either recover or enter a *D*_1_ compartment, reperesenting individuals who will eventually succumb to the disease, with probability *ρ* and after a mean time of 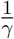. Individuals in *D*_1_ then enter the final death compartment *D*_2_ with mean time 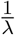. The distinction between *D*_1_ and *D*_2_ aids in accounting, and helps separate out a parameter governing the time between infectiousness and death, which is useful for model parameterization.

On a given day *t*, the following differential equations describe the instantaneuous changes in each compartment with respect to the continuous time index *τ* ∈ (*t, t* + 1]:

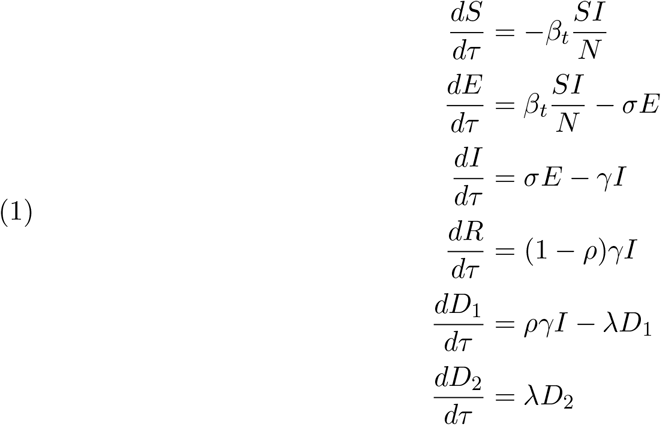

In addition, we augment the dynamics with an extra variable *C*(*τ*) to count the cumulative number of individuals that enter the *I* compartment, with dynamics 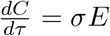 that capture only the flow into, and not the flow out of, *I*. The number of individuals that first become infectious on day *t* is then *C*(*t* + 1) − *C*(*t*); we consider these individuals candidates for being detected as confirmed cases on day *t*. We do not attempt to model testing delays, or mismatches between onset of infectiousness and onset of a detectable infection.

The state vector at time *t* is then:

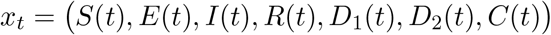

The distribution *p*(*x*_1_) of the initial state *x*_1_ is described in Appendix A2. The update from time *t* to time *t* + 1 is obtained by numerically solving the ordinary differential equation (ODE) with dynamics 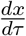 given by Equation (1) for the time interval *τ* ∈ (*t, t* + 1], over which the dynamics are fixed. We write this as:

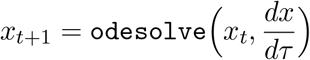

We use the ODE solver in the the Python library JAX [10], which uses the Dormand-Prince algorithm [37], a member of the Runge-Kutta family of ODE solvers. Importantly, JAX also supports automatic differentiation of odesolve using the adjoint method [12] to compute partial derivatives of *x*_*t*+1_ with respect to both the initial value *x*_*t*_ and all dynamics parameters affecting 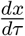. This is a key functionality that allows us to embed ODE dynamics within a probabilistic model for which NumPyro can perform inference using Hamiltonian Monte Carlo [11].

In 2020, significant efforts to control the spread of COVID-19 relied on non-pharmaceutical interventions. These included mandatory distance between individuals, closures of public spaces, and mask wearing. To add to the complexity, these interventions were implemented and repealed at different time points throughout the year, and the public complied with the interventions to varying degrees [38]. In order to capture the aggregate effect of the interventions and other behavior changes non-parametrically, we choose a flexible model for the time-varying transmission parameter. We allow *β*_*t*_ to vary following a Gaussian random walk on logarithmic scale, that is:

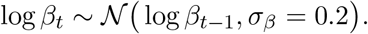

The random walk models non-stationary dynamics within the *observed* time period (*t* from 1 to *T*). For forecasting (*t > T*), MechBayes does not attempt to model future behavior changes, and simply predicts that the final value of *β*_*t*_ will persist in to the future. However, to avoid extreme senstivity of forecasts to one or a few data points near the end of the time series, we average over the last 10 days instead of taking the final value, that is, for all *i* ≥ 1:

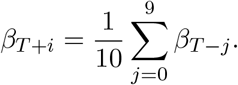

#### 2.3.2. Observation Model

The observed data used to fit the model is based on time series of incident confirmed cases and deaths. The model is fit separately for each location. The observations on day *t* are *y*_*t*_ = (*y*_*t,c*_, *y*_*t,d*_), where *y*_*t,c*_ is the number of new cases confirmed on day *t*, and *y*_*t,d*_ is the number of new reported deaths. We assume that *y*_*t,c*_ is a noisy observation of *C*(*t* + 1) − *C*(*t*), the modeled number of new infections on day *t*, using the NB2 negative binomial model for for overdispersed counts [39]:

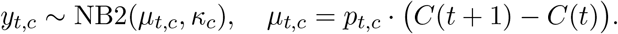

This satisfies 𝔼[*y*_*t,c*_] = *µ*_*t,c*_, with the parameter *p*_*t,c*_ acting as a detection rate on the modeled number of new infections; the parameter *κ*_*c*_ controls overdispersion, with 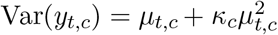. Note that the detection rate *p*_*t,c*_ is time-varying (see below). Similarly, we assume that *y*_*t,d*_ is a noisy observation of *D*_2_(*t* + 1) − *D*_2_(*t*):

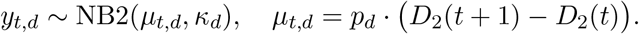

The detection rate *p*_*d*_ for deaths is not time-varying. The dispersion parameters *κ*_*c*_ and *κ*_*d*_ for both cases and deaths are estimated and given a truncated normal prior distribution with location 0.30, scale 0.15, and lower truncation limit 0.10.

We model the time-varying case detection rate as:

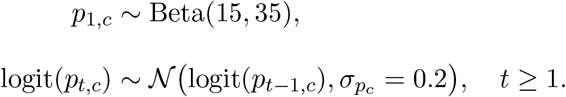

The Beta distribution on the case detection rate at time *t* = 1 (corresponding to early March in our operational model) satisfies 𝔼[*p*_*c*_] = 0.3, with 90% probability of falling between 0.22 and 0.38. Preliminary experiments suggested that the detection rate is poorly determined by data and short-term forecasts are not sensitive to this parameter.^2^ We therefore use a moderately strong prior centered at 30%, as suggested by the literature [40]. We then allow the detection rate to vary over time following a Gaussian random walk on the logodds scale, as shown above. This is meant to loosely model changes in diagnostic testing over time; in practice, it provides flexibility in the model that likely captures other changes in the relationship between reported cases and deaths over time, such as changes in the fatality ratio of the population infected at a given time.

For deaths, we place a strong prior on the reporting rate: *p*_*d*_ ∼ Beta(90, 10). This satisfies 𝔼[*p*_*d*_] = 0.9 with 90% probability between 0.89 and 0.92. That is, we assume that deaths due to COVID-19 are most often correctly reported [41]. As with the absolute value of the case detection rate, short-term forecasts are not very sensitive to this parameter.

#### 2.3.3. Epidemiological Model Parameters

We use relatively informative priors for epidemiological parameters, such as *γ, σ, ρ, λ*, and initial compartment values. The details are described in the Appendix A1. However, the identifiability of model parameters in compartmental models where the data consists only of a time series of incident cases and deaths presents a problem for uninformative priors [5]. MechBayes allows for a time-varying reproductive number through the random walk on *β*_*t*_, while maintaining strong priors on other epidemiological parameters.

#### 2.3.4. Implementation

The components described so far lead to the full probability model

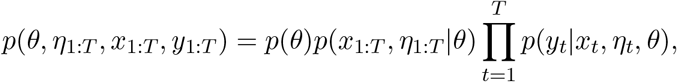

where *η*_*t*_ = (*β*_*t*_, *p*_*t,c*_) are time-varying parameters (contact-rate and case detection rate) and

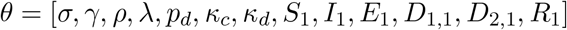

is the vector containing all other parameters.

Each model component is implemented in NumPyro [9]. We then use NumPyro’s implementation of the No-U-Turn Sampler [42] (a variant of Hamiltonian Monte Carlo [11]) to draw samples from the posterior distribution *p*(*θ, η*_1:*T*_, *x*_1:*T*_ |*y*_1:*T*_) given an observation sequence *y*_1:*T*_ (for model diagnostics), and to sample from the distribution *p*(*y*_*T* +1:*T* +*k*_|*y*_1:*T*_) to make forecasts of future reported cases and deaths.

We draw 1000 warm-up sample and then 1000 posterior samples of model parameters. We also monitor the number of effective samples produced by HMC to ensure it is large enough to reflect accurate exploration of the posterior [43].

### 2.4. Operational Forecasts

On May 10, 2020, we began submitting incident and cumulative death forecasts on a weekly basis to the US Centers for Disease Control (CDC) through the COVID-19 Forecast Hub consortium [14].^3^ Each week, we submitted 1–4 week ahead forecasts for the 50 US states and Washington, D.C., and later added forecasts for the US national-level and US territories. All forecasts used daily data up to and including the Sunday before the Monday submission. The “1 week ahead” forecast corresponds to the week ending on the following Saturday, the “2 week ahead” forecast to the week ending on the second following Saturday and so on. The model remained remained stable from May 10 until the time of writing, with only minor changes, e.g., to prior distributions.

Over time, we developed a quality-assurance process to tune our model and detect and troubleshoot suspicious forecasts. We regularly monitored the performance of our recent forecasts in terms of mean absolute error and calibration of prediction intervals as measured by the probability integral transform [44]. We used these metrics and diagnostic plots to compare submitted forecasts to alternate models to tune parameters. This led us to introduce a resampling procedure to mitigate too-large prediction intervals (on May 17, 2020) and to slight changes to prior distributions (on September 6 and October 20, 2020).

Suspicious forecast were primarily caused by data reporting issues. It was relatively common for a state to report a large backlog of cases or deaths on one day due to changes in reporting practices or to correct previous errors. As an extreme example, New Jersey (NJ) reported nearly 1,600 daily deaths on June 25, 2020 when it began the practice of including deaths from probable COVID-19 cases in its totals. Similarly, Texas (TX) removed 3,000 confirmed cases on July 7, 2020 when it determined that cases detected by antigen testing should not be reported. Changes of smaller magnitude were extremely common. Because MechBayes includes a flexible model of time-varying transmission, it interprets large changes in cases or deaths as evidence of significantly increased or decreased transmission, which leads to unrealistic forecasts.

Our quality-assurance process involved viewing diagnostic plots of each forecast together with the recent time series of incident deaths and cases to identify forecasts that were unduly influenced by data reporting issues. We also checked the JHU CSSE website [15] for notifications of reporting issues that might not be obvious in diagnostic plots. After identifying a potential problem, we searched for documented evidence of a reporting issue. These were usually reported on state department of health websites or by local news outlets. If we could identify a reporting issue, we distributed the excess number of incident cases or deaths evenly over some time window selected using our best judgment based on available information.

We made a small number of other interventions. Some states do not report data on Saturdays or Sundays; we modified the data to omit such observations instead of treating them as zeros. Starting in October, we sometimes omitted weekend observations even if they were nonzero to mitigate the influence of low values that are due only to the weekly reporting cycle. In a small handful of cases, the inference routine failed to converge or diagnostics showed signs of numerical instability; in those cases, we adjusted the prior distributions slightly and reran the model to overcome the problem.

### 2.5. Experimental Setup

We conducted two different evaluations. First, we evaluated the forecasts made in real time and submitted to the CDC via the COVID-19 Forecast Hub to assess the quality of MechBayes as an operational forecast model. Second, we conducted an ablation study that compared retrospective forecasts made using different versions of MechBayes to assess the importance of different model components on forecast accuracy.

#### 2.5.1. Baseline Forecast Evaluation

We evaluated all 1–4 week ahead incident death forecasts submitted to the CDC between May 10, 2020 and October 18, 2020 for the 50 US states and Washington, D.C.^4^ We computed the absolute error (AE) for point forecast and examined the distributions of absolute errors for different locations, forecast horizons, and dates. We used mean absolute error (MAE) as a summary metric. In addition, to evaluate the uncertainty calibration of our probabilistic forecasts, we measured the empirical coverage rates of the prediction intervals obtained from the forecasted quantiles by measuring the fraction of actual observed values that fell within different prediction intervals.

We compared the performance of MechBayes to the performance of the COVID-19 Forecast Hub baseline model described by Ray et. al [1] and used a random effects regression model to assess the statistical significance of absolute error differences between the two models; see Appendix 6.1.

#### 2.5.2. Forecast Hub Alternate Model Comparison

To evaluate the relative performance of MechBayes against other models submitted to the Forecast Hub, we chose the 10 models (including MechBayes) that have been submitting forecasts from June 21, 2020 to October 18 2020 for incident deaths across all 50 states and D.C.. These criteria balance including as many models as possible, including ones that have performed well in other analyses, while also having a large number of locations and dates for which all of the models made forecasts. For each of the models, we examined the distribution of absolute error of all point forecasts, as well as summary metrics such as the mean and median absolute error. We include this analysis to demonstrate that for a particular common subset of locations and dates, MechBayes is a top performing model. A comprehensive evaluation of Forecast Hub models by Cramer et al [45] examines multiple performance metrics and addresses the problem of comparing models that make forecasts for different sets of locations and dates, and also finds that MechBayes is one of the top two models among those submitting forecasts from May 17, 2020 to October 26, 2020.

#### 2.5.3. Ablation Test

We also performed a retrospective evaluation to demonstrate the improvement in accuracy due to addition of different model components. We define the following three variants of MechBayes:

- MechBayes Full is the full MechBayes model as submitted to the Forecast Hub and described in the previous sections, including observations of both reported cases and deaths and a time-varying random-walk model for the case detection rate *p*_*t,c*_.
- MechBayes Fixed-Detection is the same as MechBayes full but with the time-varying detection rate *p*_*t,c*_ replaced by a constant detection rate *p*_*c*_.
- MechBayes Fixed-Detection Death-Only is the same as MechBayes Fixed-Detection but with observations only on incident deaths (the forecasted quantity), and not on cases. This model is included to assess the value of using incident cases as evidence to help forecast incident deaths.

Other than the changes described above, all model components, data handling, and fitting procedures were identical. Note that we did not include a model without time-varying transmissibility. Such a model is unable to adequately fit the observed data; previous COVID-19 modeling has clearly established that time-varying transmissibility is an essential model component [6, 28, 33, 46].

## 3. Results

### 3.1. Model Fitting and Inference

MechBayes captures signal in the observed data, even in the presence of highly variable incident death reporting, and produces forecasts and prediction intervals that track the data well (Fig. 2A). The model infers a relationship between the logarithm of incident deaths and time that is nearly linear over short time periods but with slopes that change over time at somewhat discrete time points (Fig. 2B), highlighting the exponential growth (or decline) over short time periods that is a hallmark of compartmental models, but also the fact that these dynamics vary over longer time periods.

**Figure 2.**
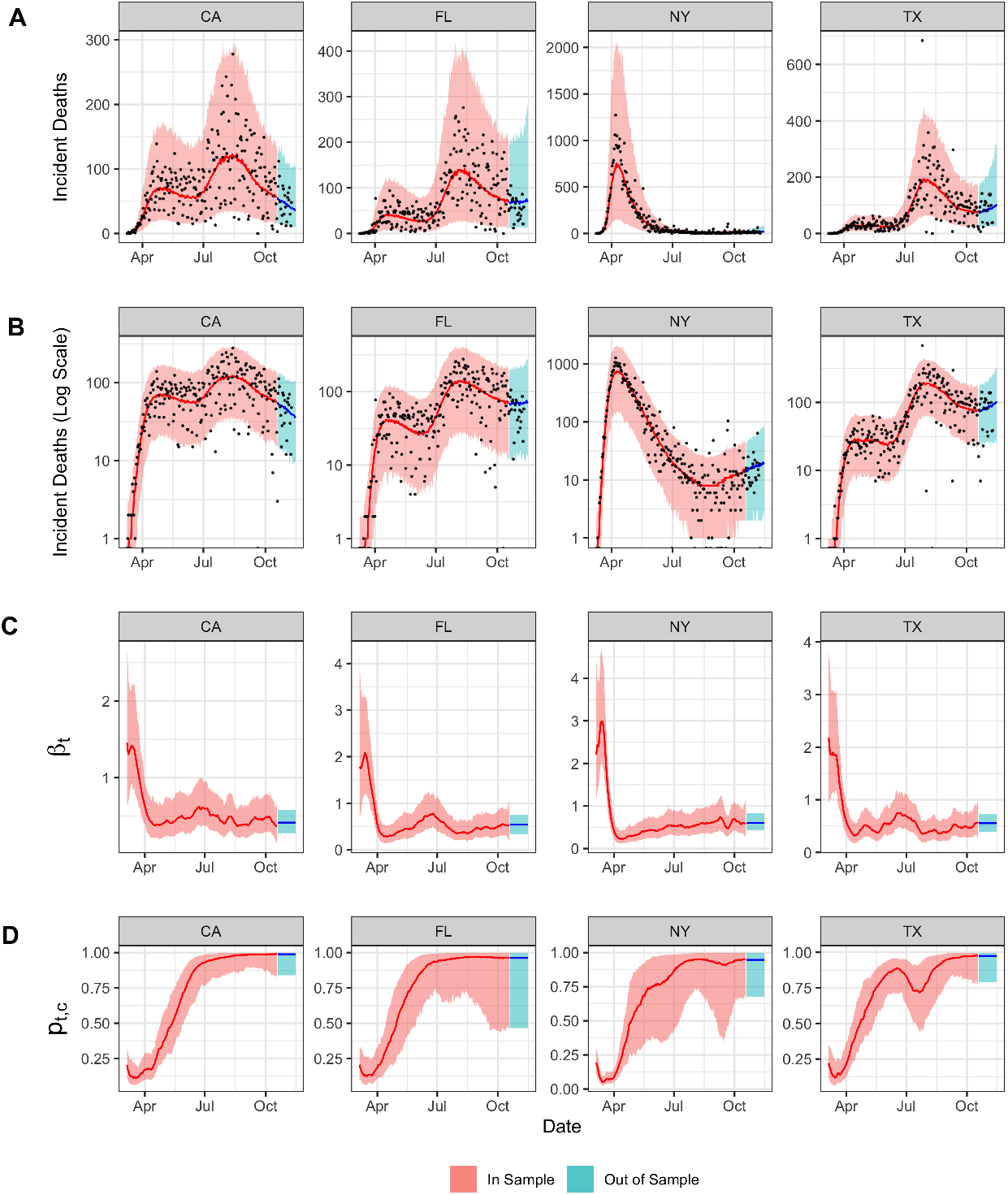
**A, B**. Example posterior fits as well as 1–4 week ahead forecasts made on October 18, 2020 for four selected states. Shaded regions show 95% prediction intervals for in-sample (red) and forecast (blue) posterior predictive distributions; lines show posterior medians; points show observed data. **C**. Posterior median and 95% credible interval of time-varying contact rate *β*_*t*_ for each of the four states. **D**. Posterior median and 95% credible interval of the time-varying ratio between cases and deaths parameter (*p*_*t,c*_).

The inferred value of the time-varying contact rate parameter *β*_*t*_ (Fig. 2C) is closely tied to the observed rate of change of incident deaths (and cases, not shown), especially as observed on a logarithmic scale: *β*_*t*_ is high across all four example states in mid-March when incident deaths grew rapidly, then falls as the growth rate of incident deaths declines during and after the initial wave, with subsequent changes that can be matched to specific events in the states, e.g., increases in *β*_*t*_ associated with second waves in Texas, California, and Florida during the summer, and a slow increase in *β*_*t*_ in New York associated with an eventual increase in deaths in the fall. In all four states, the inferred value of time-varying case detection rate *p*_*t,c*_ increases significantly from the start of the pandemic (Fig. 2D). In practice, this parameter likely functions to model *any* changes over time in the ratio of observed cases to observed deaths. One reason for such a change is increased diagnostic testing; another reason is a decrease in the overall infection-fatality ratio (e.g., due to changes in the age distribution of patients and improved treatments). Both changes would lead to a larger number of observed cases for the same number of deaths, and likely occurred in conjunction, leading the model to significantly increase its estimate of *p*_*t,c*_ over time. It is apparent that MechBayes also uses *p*_*t,c*_ to absorb some reporting anomalies, as seen in Texas: a string of both abnormally high and low numbers of incident deaths were reported in late summer, which correspond to the model inferring a temporary decrease in *pt,c*.

### 3.2. Real-time Forecast Results

#### 3.2.1. Baseline Comparison

MechBayes had lower absolute error than the baseline model in all quantiles of the error distribution except for the maximum (Fig. 3A). The gap in performance (as measured by absolute error) increased as the magnitude of the error increased up until the 0.99 quantile of the error distribution. Above the 0.99 quantile of error distribution MechBayes still performed better for all but one quantile, however, the gap in performance closed. MechBayes also had a lower absolute error at the central tendency of the absolute error distribution (as measured by mean or median) (Fig. 3B).

**Figure 3.**
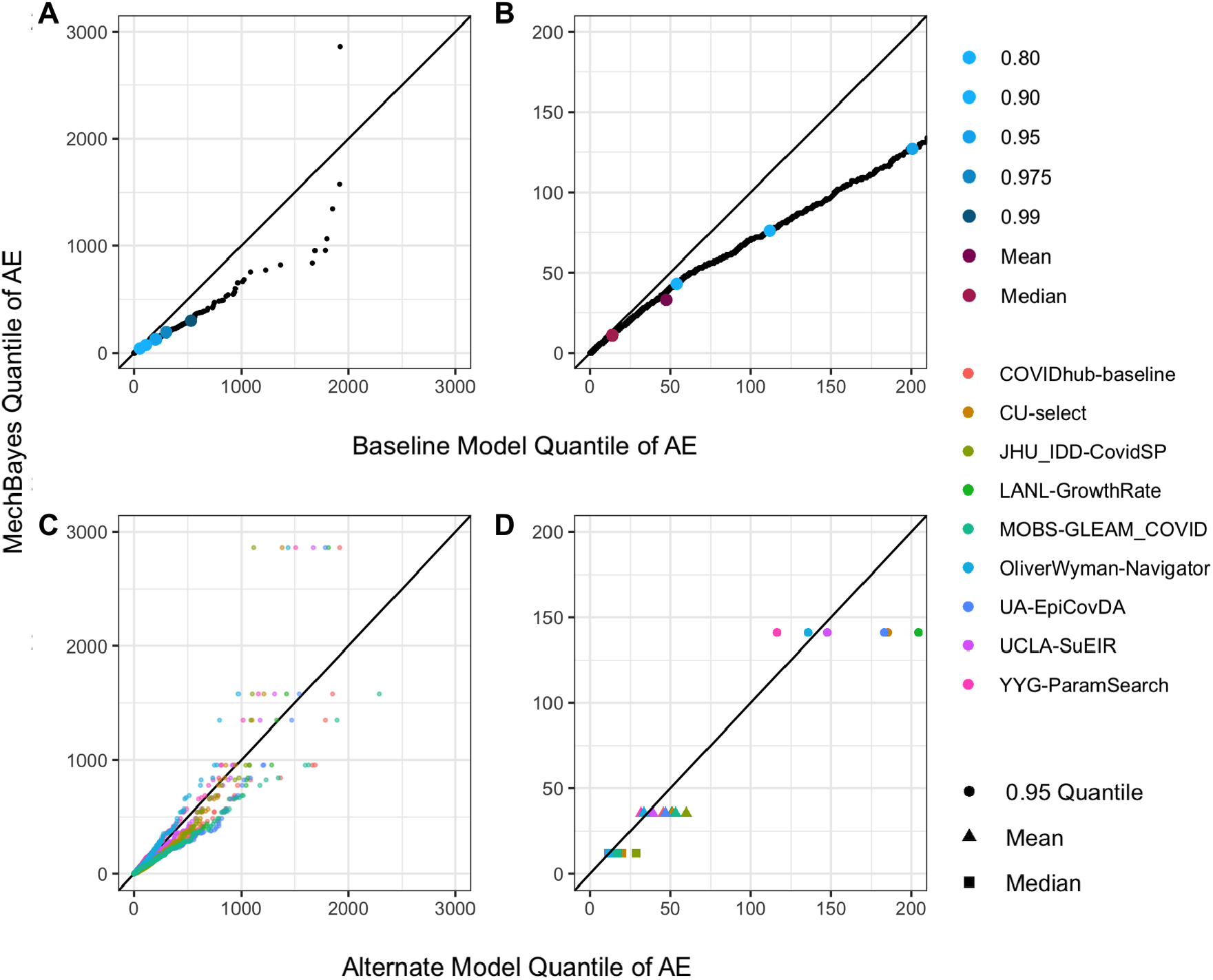
**A** Quantile-quantile plot of absolute error distribution for MechBayes (y-axis) vs. baseline model (x-axis) over every combination of location, forecast date, and target, along with summary metrics. MechBayes absolute error is lower than the baseline error at every quantile except the largest. **B** Quantile-quantile plot of absolute error distribution zoomed in to errors less than 200 deaths. There are more significant improvements relative to the baseline when the baseline error is high (e.g. more than 60). **C** Quantile-quantile plot of absolute error distribution for MechBayes (y-axis) vs. alternate models (x-axis) submitted to the Forecast Hub. Each point represents the absolute error for a combination of location, forecast date, and target. **D** Mean, median and 0.95 quantile of the absolute error distribution for MechBayes (y-axis) and alternate models (x-axis). MechBayes median and mean of the absolute error distribution is lower for all but one model.

Overall, MechBayes had an MAE of 32.85 deaths, when averaging over all regions, forecast dates, and targets. The baseline model had an MAE of 47.06 deaths. The prediction intervals at the 95% level covered the truth 94.8% of the time for MechBayes, compared to 89.2% for the baseline model over all targets, regions, and forecast dates.

MechBayes had similar or lower MAE than the baseline for most states and targets (Figs. 4A,4C). In particular, for the locations with the highest total death counts (NY, TX, CA, FL) [15], MechBayes uniformly outperformed the baseline with the only exception being Florida (FL) for the 4 week ahead target. When absolute errors were small, both models performed similarly.

**Figure 4.**
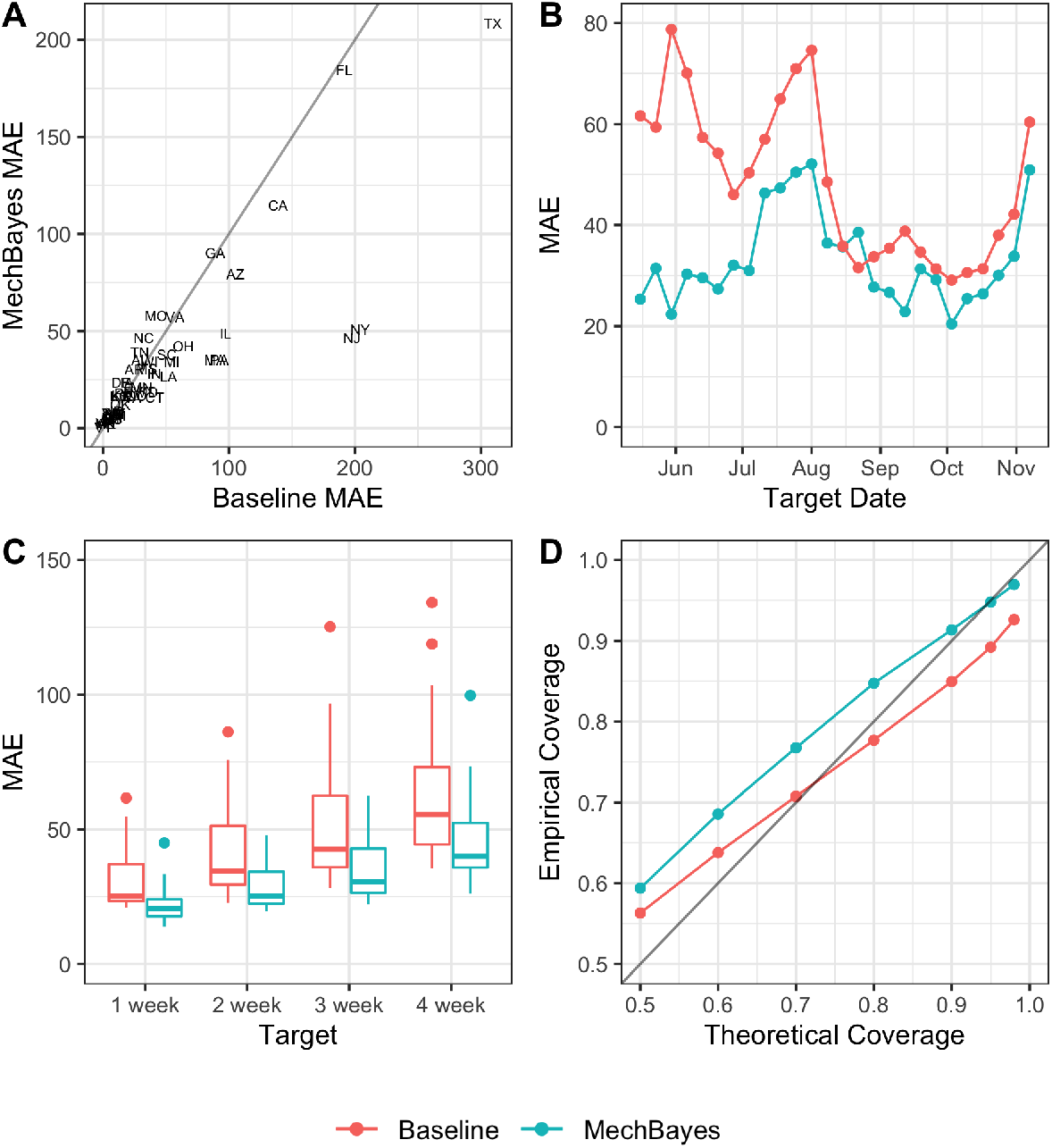
**A** Mean absolute errors for MechBayes and the baseline model averaged over all forecast dates and targets for each location. Notice that for states with the largest number of deaths, New Jersey (NJ), New York (NY), Florida (FL), Texas (TX), California (CA), MechBayes uniformly outperforms the baseline. **B** Mean absolute error box plots for MechBayes and baseline model by target. Each box plot shows the distribution of MAE values for all forecast dates, where one data point is the MAE over all locations for a single date. MechBayes has lower quartiles of mean absolute error across all targets. **C** Mean absolute errors for MechBayes and the baseline model averaged over all regions and targets by target end date: a point on panel B represents the absolute error of the 1–4 week ahead forecast made for that date. MechBayes has lower mean absolute error for 21 of the 23 forecast dates. **D** Percent of observations (y-axis) falling within the prediction interval at the given level of confidence (x-axis) for both MechBayes and the baseline model. MechBayes intervals are better calibrated than the baseline at high confidence levels and slightly too wide at lower confidence levels.

MechBayes improved uniformly over the baseline model for every target (Fig. 4C); p *<* 0.01, see Appendix 6.1), with MechBayes MAE ranging from 21.73–45.48 for 1–4 week ahead and baseline model MAE ranging from 30.60–63.01 for 1–4 week ahead, averaged over all forecast dates. The mean absolute error increased as horizon increased, which is to be expected. The distribution of errors by forecast date for a given target showed significant variability, suggesting that different targets were easier to predict on certain forecast dates, again reflecting the change in forecast difficulty throughout epidemic.

MechBayes had lower MAE by forecast date (averaged over locations and targets) than the baseline for 21 out of 23 forecast weeks (Fig. 4B). The largest increase in incident deaths during the evaluation period (May 11, 2020 through October 18, 2020) occurred in early July 2020. MechBayes significantly outperformed the baseline model during these weeks. However, in weeks with a small increase or a decrease in incident deaths, the MAEs were much closer. This suggests again that MechBayes performs well during periods of more rapid change in incident deaths.

MechBayes prediction intervals contained the truth with at least the predicted probability (Fig. 4D), but were somewhat conservative: the empirical probability of containing the truth was nearly exact for the 95% interval, and higher than predicted for smaller intervals.

#### 3.2.2. Alternate Model Comparison

MechBayes had a lower absolute error in nearly all quantiles of the error distribution for 8 out of the 9 alternate models submitted to the Forecast Hub (Fig. 3C). MechBayes was in the top 3 out of the 10 models based on mean, median, and the 0.95 quantile of the absolute error distribution (Fig. 3D). Specifically, YYG-ParamSearch (lowest MAE) had an MAE of 31.42, OliverWyman-Navigator had an MAE of 33.41 and MechBayes had an MAE of 35.40. MechBayes performed worse relative to other models for its top three error values, and had the second-largest maximum error out of the 10 models.

The median of the error distribution for all models was clustered around 15 deaths, with little separation between models. The mean absolute error was clustered around 40 deaths with slightly more separation between models. The 0.95 quantile of the error distribution at around 175 deaths began to show more significant separation between models (Fig. 3D).

### 3.3. Ablation Test Results

MechBayes Full produced consistently better point forecasts than MechBayes Fixed-Detection Death-Only or MechBayes Fixed-Detection (Fig. 5A). When averaged over all targets, locations and forecast dates MechBayes Full had an MAE of 27.85, MechBayes Fixed-Detection Death-Only had an MAE of 37.5, and MechBayes Fixed-Detection had an MAE of 134.69. At every quantile level, the error of MechBayes Fixed-Detection was significantly larger than that of MechBayes Fixed-Detection Death-Only, which suggests that using deaths as evidence is only beneficial in conjunction with flexibility allowed by the time-varying ratio between cases and deaths (*p*_*t,c*_). However, the interval coverage performance shows that MechBayes Fixed-Detection was closer to the nominal coverage than MechBayes Fixed-Detection Death-Only (Fig. 5B). Both MechBayes Full and MechBayes Fixed-Detection prediction intervals contained the truth with approximately the correct probability; however, MechBayes Fixed-Detection Death-Only intervals were too small and contained the truth significantly less frequently than expected.

**Figure 5.**
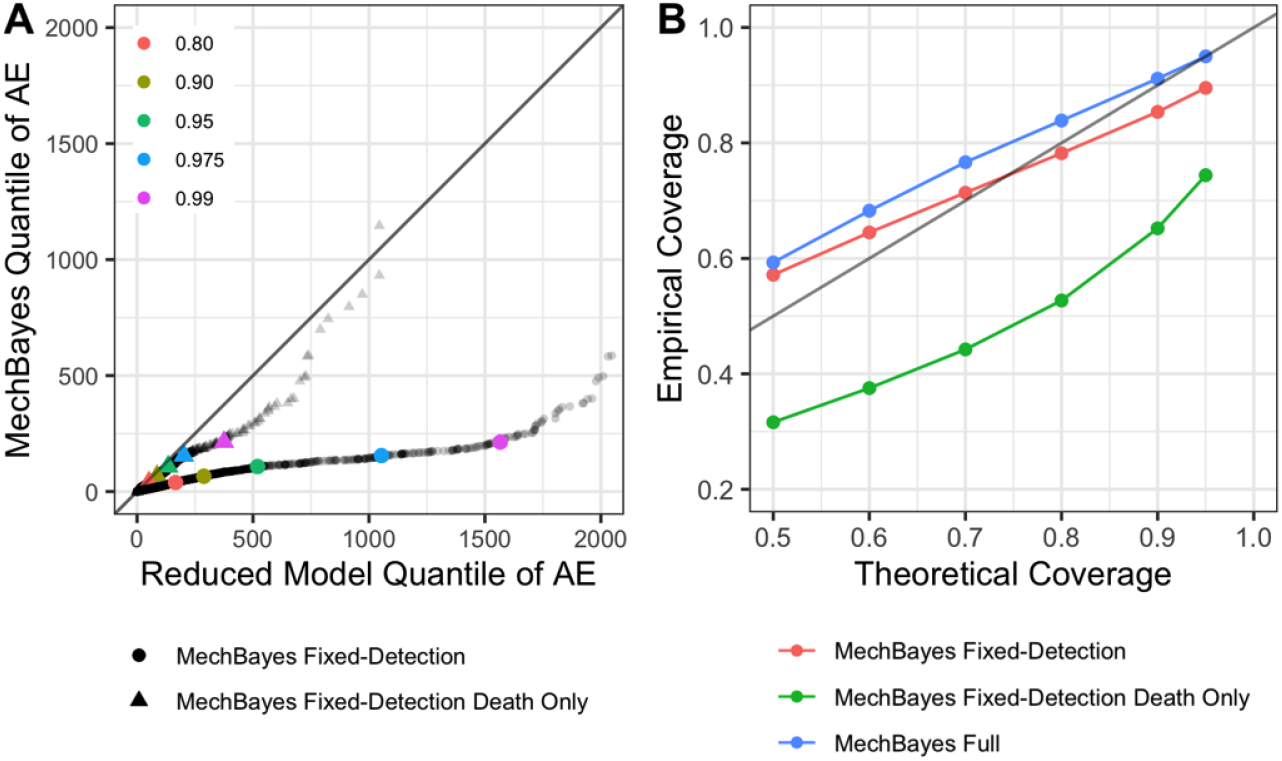
**A** Absolute error quantiles of MechBayes Full (y-axis) against the reduced models, MechBayes Fixed-Detection Death-Only and MechBayes Fixed-Detection. MechBayes Full uniformly improves over MechBayes Fixed-Detection and improves in all but the maximum quantile over MechBayes Fixed-Detection Death-Only. **B** Percent of observations (y-axis) falling within the prediction interval at the given confidence level (x-axis). MechBayes Fixed Detection seems to be closest to the nominal level of coverage, suggesting that adding the uncertainty in the ratio between observed cases and observed deaths made the model slightly under-confident. In contrast, using only observations on deaths significantly compromised model uncertainty.

## 4. Discussion

MechBayes is a fully Bayesian compartmental model capable of accounting for varying transmission rates, observations on both cases and deaths, and a time-varying ratio of cases to deaths. MechBayes produced consistently accurate real-time forecasts over the course of 23 evaluation weeks, and was ranked as one of the top 2 of 10 models on median and mean absolute error. Our experiments led us to the following conclusions about the performance of this model and the underlying methodology.

- **MechBayes is accurate when compared to a baseline model**. MechBayes had lower absolute error than the baseline model in almost all quantiles of the error distribution (Fig. 3). The performance gain was higher when predicting deaths was difficult (in the upper quantiles of the absolute error distribution) because deaths were changing rapidly. This is true across target, week, and region breakdown. Error is significantly lower for 1–4 week ahead predictions, with larger improvements at longer horizons. Additionally, the biggest gains in performance occur in regions with the largest incident death counts (Fig. 4A), such as Texas (TX), California (CA), New York (NY), New Jersey (NJ) and Florida (FL) [15]. Finally, MechBayes performance gain was highest in forecast weeks with the large absolute error (Fig. 4B). This leads us to conclude that MechBayes is better than the baseline model when it really counts: in regions where deaths were high and in weeks that were difficult to predict because of rapidly changing incident deaths.
- **MechBayes is accurate when compared to the alternate models submitted to the Forecast Hub**. MechBayes ranked second out of eleven models in terms of mean and median of the absolute error distribution (Fig. 3D). While MechBayes is one of the top performing models submitted to the Forecast Hub, its largest errors are higher than alternate models (Fig. 3D). The same mechansims— the underlying exponential growth intrinsic to compartmental models and the flexible, time-varying transmission—that allow MechBayes to accurately model the pandemic in many situations also make its forecasts highly sensitive to errors estimating the current rate of exponential growth. For example, the four-week forecast for Florida (FL) on July 25, 2020 was too high by 2861 deaths due to MechBayes estimating a high exponential growth based on recent trends and possible reporting issues, when the eventual growth rate over the next four weeks was much more modest.
- **MechBayes is probabilistically well-calibrated**. The MechBayes 95% prediction interval contains the truth 94.6% of the time (Fig. 4D). MechBayes is conservative for smaller intervals. As a Bayesian model, MechBayes is able to reason effectively about uncertainty in the epidemiological model parameters, state variables, and observation noise given the evidence and translate this into appropriately calibrated forecast uncertainty.
- **Adding case data when predicting deaths is helpful but only when accounting for time-varying relationship between observed cases and deaths**. Allowing for a time-varying ratio between cases and deaths is a key feature for lower MAE (Fig. 5A). MechBayes Full both incorporates incident cases as evidence and allows for a flexible deviation between cases and deaths, which makes the model consistently more accurate than a model that does not account for cases at all (MechBayes Fixed-Detection Death-Only) and a model that does account for cases but fixes the detection probability (MechBayes Fixed-Detection). Including cases without properly accounting for factors that yield a changing ratio between observed cases and observed deaths over time hurts performance over leaving out observations on cases all together.

We have seen that MechBayes is a powerful Bayesian compartmental model that can capture the real-world complexities of forecasting during a pandemic. MechBayes’ disease model is a classical compartmental model, which has good inductive bias for a novel epidemic. MechBayes is fully Bayesian, which allows for a balance between model structure, evidence through observations on cases and deaths, and uncertainty. The implementation in the NumPyro probabilistic programming language allowed for rapid model development and experimentation. Finally, a reasonable investment of effort in validation prevented model pathologies due to data quality issues.

While we chose an exponential random walk on *β*_*t*_ there are many other choices for flexible non-parametric modeling of transmissibility. Further work might consider a spline model, Gaussian process, or a semi-parametric model capable of taking intervention dates as covariates. Additionally, as more COVID-19 data streams come online, more observation models on compartments can be added to MechBayes and fit using the same framework. Other methods of expressing compartmental models (e.g. the renewal framework in [33]) may lead to more efficient and flexible implementations. Modeling more characteristics of the surveillance system (such as weekly reporting) may also improve forecast performance. Through real-time and retrospective evaluation, we demonstrated the success of MechBayes in forecasting COVID-19 both in terms of point and probabilistic forecasts. The model is able to improve over the baseline model as well as reduced forms of MechBayes, and is ranked in the top two models out of the 10 considered that submitted forecasts to the Forecast Hub since May, 2020. While future pandemics may be unavoidable, MechBayes is a flexible framework that can be adapted to the unique challenges of pandemic forecasting efforts.

## Data Availability

The manuscript uses the COVID-19 incident case and death data provided by JHU CSSE.

https://github.com/CSSEGISandData/COVID-19

## 5. Acknowledgments

We would like to thank Evan Ray for many productive conversations during the development of MechBayes. This material is based upon work supported by the National Science Foundation under Grant No. 1749854. This work has been supported by the National Institutes of General Medical Sciences (R35GM119582). The content is solely the responsibility of the authors and does not necessarily represent the official views of NIGMS or the National Institutes of Health.

## 6. Appendix

### 6.1. Statistical Test for Significance

To assess the significance of absolute error differences between MechBayes and the baseline model we use a random effects regression model of the form,

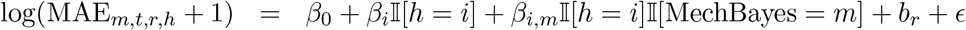

where 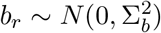, *ϵ* ∼ *N* (0, *σ*^2^), *h* represents the 1–4 week ahead target (horizon), *m* is an indicator for MechBayes, *t* is timezero, and *r* is region. We chose this model because it explains the variation in MAE by model and horizon while allowing varying baseline MAE values by region. Here, variation over time in MAE within a specific region is explained by differences in model performance This leads to the following coefficient estimates for the fixed effects using the lme4 package [47].

**Table 1.** Coefficient estimates and t-values for MAE evaluation model. We can see that MechBayes performs statistically significantly better than the CDC baseline model for 1-4 weeks ahead.

|  | Estimate | Std. Error | df | t value | Pr(>\|t\|) |
| --- | --- | --- | --- | --- | --- |
| $\beta_0$ | 2.79 | 0.14 | 55.27 | 19.79 | 0 |
| $\beta_1$ | -0.34 | 0.04 | 9389 | -8.16 | 0 |
| $\beta_2$ | -0.09 | 0.04 | 9389 | -2.17 | 0.03 |
| $\beta_3$ | 0.03 | 0.04 | 9389 | 0.72 | 0.47 |
| $\beta_4$ | 0.20 | 0.04 | 9389 | 4.45 | 0 |
| $\beta_{1,\text{MechBayes}}$ | -0.54 | 0.04 | 9389 | -12.67 | 0 |
| $\beta_{2,\text{MechBayes}}$ | -0.31 | 0.04 | 9389 | -7.34 | 0 |
| $\beta_{4,\text{MechBayes}}$ | -0.15 | 0.04 | 9389 | -3.496 | 0 |

### 6.2. Seeding Epidemic

Due to the under-reporting of cases, we cannot use the observed data to seed the epidemic. We instead allow the model to find the initial state values for all compartments except the number of susceptible people, which we take as the population size of the geographic region minus the sum of the initial values for the other compartments to enforce the constraint that total number of individuals in all compartments is equal to the population size. We do this by assigning uniform probability to all initial states where the number of people in any given compartment does not exceed 2% of the total population. This is a highly conservative estimate for the number of infected, exposed, dead and recovered people at the start of the epidemic which is most likely much lower than 2% of the population.

This leads to the follow distributions for the initial number of individuals in each compartment, where we use the convention that the starting time is *t* = 1 to match the conventions of the discrete-time model in which these differential equations are embedded:

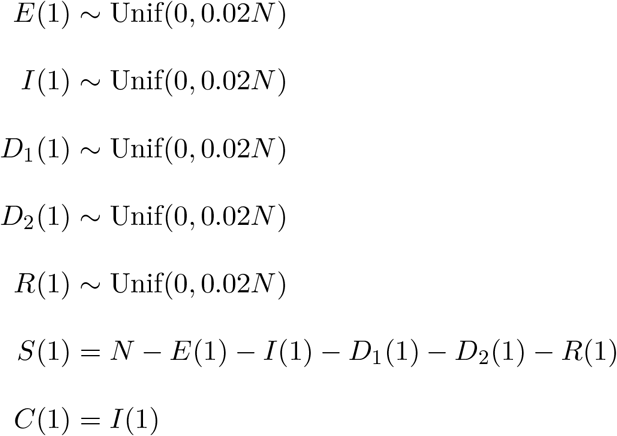

The initial state vector is then:

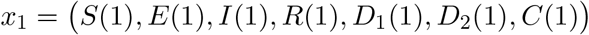

### 6.3. Priors

We also place the following priors on the transition parameters:

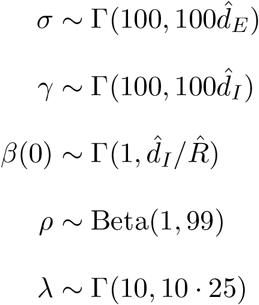

Preliminary experiments showed that the model is not highly sensitive to most of these settings, which is consistent with the non-identifiablility of compartmental models from case and death surveillance data [5], which can be viewed as overparameterization relative to the available data. Our prior on rate for leaving the exposed compartment *σ* satisfies 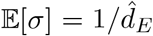, where 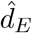 is an initial guess of the duration of the latent period. We assume 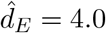 based on published estimates (shortened slightly to account for possible infectiousness prior to developing symptoms [40]. Our prior on the rate for leaving the infectious compartment *γ* satisfies 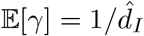, where 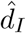 is an initial guess for the duration of infectiousness. The setting is 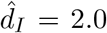 to model the likely isolation of individuals after symptom onset [48]. Our prior on the initial transmission rate is derived from the relationship between the basic reproductive number *R*_0_ and the length of the infectious period: 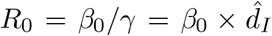. Therefore, we set our prior on the initial transmission rate to satisfy 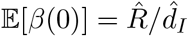 where 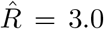 is an initial guess for *R*_0_ and 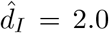, as described above. Our prior on the fatality rate *ρ* satisfies 𝔼[*ρ*] = 0.01 with 95% probability of being between [0.0002, 0.0375]. Finally, our prior on the rate *λ* at which dying patients succumb satisfies 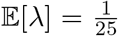 with shape 10 corresponding to roughly 25 days in the *D*_1_ compartment. Our observation model required priors on the NB2 concentration parameters for both cases and deaths, both of which were set to 0.3.

## Footnotes

1 We will also use the state variable name as a short name for the compartment itself—for example, “the *E* state”—with the correct interpretation alwaying being clear from context.

2 It primarily impacts inferences about the true number of infections in the population; forecasts are therefore expected to be more sensitive to this parameter as herd immunity is approached.

3 For two weeks prior to May 10, we submitted forecasts of cumulative deaths only while the model was under active development and lacked several of the main components described here.

4 We submitted forecasts for US territories and the US as a whole starting after May 10, but omit these from evaluation to allow for the largest number of evaluation dates where forecasts were made across a consistent set of regions.

